# The effect of a national lockdown in response to COVID-19 pandemic on the prevalence of clinical symptoms in the population

**DOI:** 10.1101/2020.04.27.20076000

**Authors:** Ayya Keshet, Amir Gavrieli, Hagai Rossman, Smadar Shilo, Tomer Meir, Tal Karady, Amit Lavon, Dmitry Kolobkov, Iris Kalka, Saar Shoer, Anastasia Godneva, Ori Cohen, Adam Kariv, Ori Hoch, Mushon Zer-Aviv, Noam Castel, Anat Ekka Zohar, Angela Irony, Benjamin Geiger, Yuval Dor, Dorit Hizi, Ran Balicer, Varda Shalev, Eran Segal

**Author notes:** Equal contribution (Ayya Keshet, Amir Gavrieli, Hagai Rossman, Smadar Shilo, Tomer Meir).

## Abstract

The vast and rapid spread of COVID-19 calls for immediate action from policy-makers, and indeed various lockdown measures were implemented in many countries. Here, we utilized nationwide surveys that assess COVID-19 associated symptoms to analyze the effect of the lockdown policy in Israel on the prevalence of clinical symptoms in the population. Daily symptom surveys were distributed online and included fever, respiratory symptoms, gastrointestinal symptoms, anosmia and Ageusia. A total of 1,456,461 survey responses were analyzed. We defined a single measure of symptoms, Symptoms Average (SA), as the mean number of symptoms reported by responders. Data were collected between March 15th to May 11th, 2020. Notably, following severe lockdown measures, we found that between March 15th and April 20th, SA sharply declined by 83.8%, as did every individual symptom, including the most common symptoms reported by our responders, cough and rhinorrhea and\or nasal congestion, which decreased by 74.1% and 69.6%, respectively. Individual symptoms exhibit differences in reduction dynamics, suggesting differences in the medical conditions that they represent or in the nature of the symptoms themselves. The reduction in symptoms was observed in all the cities in Israel, and in several stratifications of demographic characteristics. Between April 20th and May 11th, following several subsequent lockdown relief measures, the decrease in SA and individual symptoms halted and they remain relatively stable with no significant change. Overall, these results demonstrate a profound decrease in a variety of clinical symptoms following the implementation of a lockdown in Israel. As our survey symptoms are not specific to COVID-19 infection, this effect likely represents an overall nationwide reduction in the prevalence of infectious diseases, including COVID-19. This quantification may be of major interest for COVID-19 pandemic, as many countries consider implementation of lockdown strategies.

## Introduction

Since its isolation in December 2019, the novel coronavirus COVID-19 has spread to almost every country in the world. In Israel, the first infection of COVID-19 was confirmed on February 21st 2020, and in response, the Israeli Ministry of Health (MOH) employed a series of steps in an attempt to mitigate the spread of the virus in Israel. These steps were gradually aggravated; on March 9th all Israelis returning from abroad were instructed to begin a 14-day home-isolation period upon their arrival; on March 11th gatherings were limited to a maximum of 100 people; on March 12th educational institutions were closed; on March 25th citizens are instructed to stay home, and may leave only for permitted activities within a distance of 100 meters; and on March 31th the above regulations were further exacerbated forbidding any gatherings of people from different households. Policy changes in Israel thus far were enforced on the entire Israeli population. In several cities with high infection rates, stricter regulations were implemented. April imposed new challenges for the Israeli population and the government, given the Jewish holiday Passover which is traditionally celebrated with family. To prevent gatherings during the holiday period, between April 8th and 15th, harsher restrictions were imposed, with a national quarantine declared on the eve of Passover. On April 19th, a few of the restrictions were lifted and more employees were allowed to return to work and limited social events such as prayers and weddings were allowed to take place under severe restrictions on the numbers of people. In the days that followed, further relief regulations were approved, including partial reopening of schools on May 3rd.

Policy-makers in Israel, as well as others worldwide, struggle to find the most suitable policy which will slow the rate of infection while maintaining the functionality of the economy. In the absence of a vaccine against COVID-19, control of disease spreading through large-scale physical distancing measures appears to be an effective means of mitigation ^1^. These may include closures of workplaces, schools, and a general lockdown. While many countries have implemented different policies and various degrees of lockdowns ^1–3^, the effect of these measures on the prevalence of clinical symptoms in the general population has not been quantified to date. Here, we utilize data obtained from nationwide, daily, one-minute surveys, deployed by us from the early stages of COVID-19 spread in Israel ^4^, to analyze the effect of actions implemented in Israel on the prevalence of different clinical symptoms and at high geographical resolution.

## Methods

### Data

To obtain real-time information on symptoms across the entire Israeli population, we developed a simple one-minute online questionnaire ^4^. The survey was posted online (https://coronaisrael.org/) on March 14th, and participants were asked to fill it on a daily basis and separately for each family member, including those who are unable to fill it out themselves (e.g., children and elderly). The survey is filled anonymously to maintain individual privacy and was filled out 1,456,461 times to date.

The survey includes questions on respondents’ age, gender, geographic location (city and street), isolation status, smoking habits, prior medical conditions, body temperature measurement and self-reporting of symptoms. All prior medical conditions and symptoms included in the questionnaire were meticulously chosen by medical professionals, based on symptoms which were described as prevalent in patients with COVID-19 infection (e.g. cough and fatigue) as well as symptoms which were less prevalent in these patients (e.g. nausea and vomiting) ^5,6^ to allow better discrimination of individuals with possible COVID-19 infection. Responders were also asked about the number of people (separately for adults and children) they have been in close contact with in the 24 hours prior to answering the survey. The complete questionnaire can be found in Section 1 of the Supplementary Appendix (Figure S1).

The initial symptoms included cough, fatigue, myalgia (muscle pain), shortness of breath, rhinorrhea or nasal congestion, diarrhea and nausea or vomiting. Additional symptoms, including sore throat, headache, chills, confusion and loss of taste and/or smell sensation, were added in later versions. Efforts were devoted to reaching underrepresented populations through several channels, including call centers and media appearances.

Responses from participants who did not meet reasonable criteria of reported age (0-120 years of age) and body temperature (35-43 degrees Celsius) were excluded (485 and 669, respectively). Another 18,048 responses which contained invalid values in other data fields were excluded, and another 169,306 responses recorded between March 22nd to March 26st were excluded due to infrastructure transition issues.

In order to analyze additional factors that may affect the prevalence of symptoms in the population, we utilized several additional external databases. Data on the number of COVID-19 cases in Israel per data was obtained from the Israeli MOH ^7,8^. Meteorological information on the weather in Israel during the study period was obtained from the Israel Meteorological Service ^9^. Information on mobility of individuals in Israel was obtained from the Google COVID Community Mobility Reports^10^. In order to estimate the prevalence of flu-like symptoms in the Israeli population in previous years, data from the Israeli MOH were obtained ^11^.

### Statistical analysis

To estimate the changes following the Israeli lockdown policies, we analyzed the prevalence of reported symptoms throughout the period of data collection. We analyzed the complete study population, and performed several sensitivity analyses by stratifying this population according to several demographic characteristics such as city of residence, population density in city of residence, and education level (see Section 4 in Supplementary appendix). We further employed Inverse Probability Weighting (IPW), to examine the sensitivity of our estimates to possible biases in the data (see Section 2 in Supplementary appendix). In addition, we analyzed a subpopulation that included only symptomatic individuals, which were defined by responses containing at least one reported symptom (see Section 4 in Supplementary appendix). Along with the analysis of individual symptoms, we also analyzed the average number of symptoms reported per date, which we term Symptoms Average (SA). When analyzing individual symptoms, we examined each symptom starting from the time it was included in our survey, and additionally an average of all symptoms which were incorporated in the survey at the same starting time (see Section 1 in Supplementary appendix). To capture the quantitative differences in the rate and timing of symptoms reduction, we fit a sigmoid function to each symptom as well as SA and analyze its parameters (see Section 3 in Supplementary appendix).

## Results

Overall, 1,644,969 responses were collected during the study period of March 14th and May 11th, 2020. Of these, 188,508 responses were excluded (Methods) and a total of 1,456,461 responses, 1,381,229 (94.835%) adults and 75,232 (5.165%) children were eventually included in the study. The characteristics of the responders are described in Table 1.

**Table 1.** Characteristics of study population.

| **Table 1. Characteristics of study population** |  |  |  |
| --- | --- | --- | --- |
| **Characteristic, mean (SD) or %** | **All responders**<br>**1,456,461** | **Adults**<br>**1,381,229 (94.835%)** | **Children (up to 18 years old)**<br>**75,232 (5.165%)** |
| Age (years) | 49.935 (18.568) | 52.002 (16.706) | 11.977 (5.62) |
| Sex - Male | 649,746 (44.611%) | 611,191 (44.25%) | 38,555 (51.248%) |
| Smoking (currently) | 173,666 (11.924%) | 171,248 (12.398%) | 2,418 (3.214%) |
| Currently in Isolation | 88,703 (6.09%) | 85,847 (6.215%) | 2,856 (3.796%) |
| **Symptoms** |  |  |  |
| Body temperature (Celsius) | 36.37 (0.457) | 36.364 (0.45) | 36.553 (0.633) |
| Fever (body temperature above 38 °C) | 1,551 (0.394%) | 1,214 (0.317%) | 337 (3.332%) |
| Feel good | 1,391,168 (96.445%) | 1,319,867 (96.518%) | 71,301 (95.124%) |
| Shortness of breath | 12,931 (0.888%) | 12,401 (0.898%) | 530 (0.704%) |
| Rhinorrhea or nasal congestion | 83,687 (5.746%) | 79,095 (5.726%) | 4,592 (6.104%) |
| Nausea and vomiting | 6,340 (0.48%) | 5,798 (0.462%) | 542 (0.816%) |
| Muscle pains | 19,669 (1.35%) | 19,081 (1.381%) | 588 (0.782%) |
| Sore throat | 30,658 (2.241%) | 29,453 (2.268%) | 1,205 (1.731%) |
| Fatigue | 32,042 (2.322%) | 30,429 (2.323%) | 1,613 (2.296%) |
| Diarrhea | 12,072 (0.914%) | 11,377 (0.907%) | 695 (1.046%) |
| Loss of taste or smell | 2,947 (0.214%) | 2,801 (0.214%) | 146 (0.208%) |
| Confusion | 2,512 (0.182%) | 2,312 (0.177%) | 200 (0.285%) |
| Cough | 75,255 (5.167%) | 71,375 (5.167%) | 3,880 (5.157%) |
| Dry cough | 33,306 (2.414%) | 31,647 (2.416%) | 1,659 (2.362%) |
| Moist cough | 35,317 (2.559%) | 33,731 (2.575%) | 1,586 (2.258%) |
| Headache | 29,640 (2.148%) | 28,645 (2.187%) | 995 (1.417%) |
| Chills | 5,016 (0.363%) | 4,754 (0.363%) | 262 (0.373%) |

First, we analyzed the general effect that the restrictions implemented by the Israeli MOH had on mobility patterns and social distancing in the Israeli population. As expected, while physical distancing policies were implemented, the self-reported number of children and adults with whom a person has been in close contact with (defined as within approximately 2 meters for more than 15 minutes) during the last 24 hours was stable and low. A rise in these numbers was observed once restrictions began to release, on April 20th. First, a rise in the number of self-reported adults, starting on April 20th, when more workplaces were opened and small social events were allowed to take place, followed by an increase in the number of self-reported children which rose starting from May 3rd, when schools partially reopened (Figure 1D, Table 2). A similar trend was observed in data on the mobility of the Israeli population^10^, demonstrating an increase in both workplace and recreation mobility as the restrictions were gradually lifted (Figure 1F, Table 2).

**Table 2:**
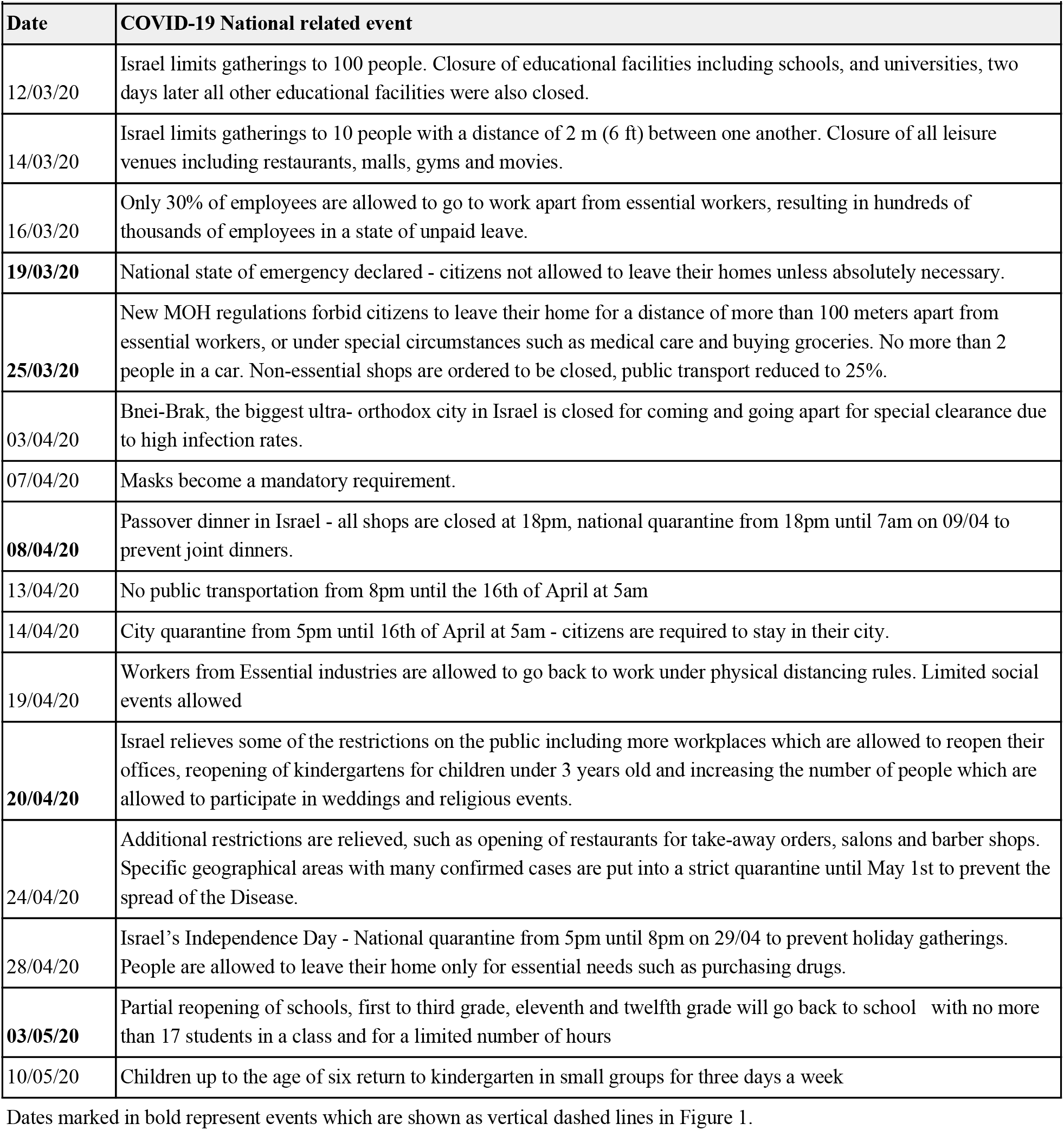
COVID-19 related national events by date.

**Figure 1:**
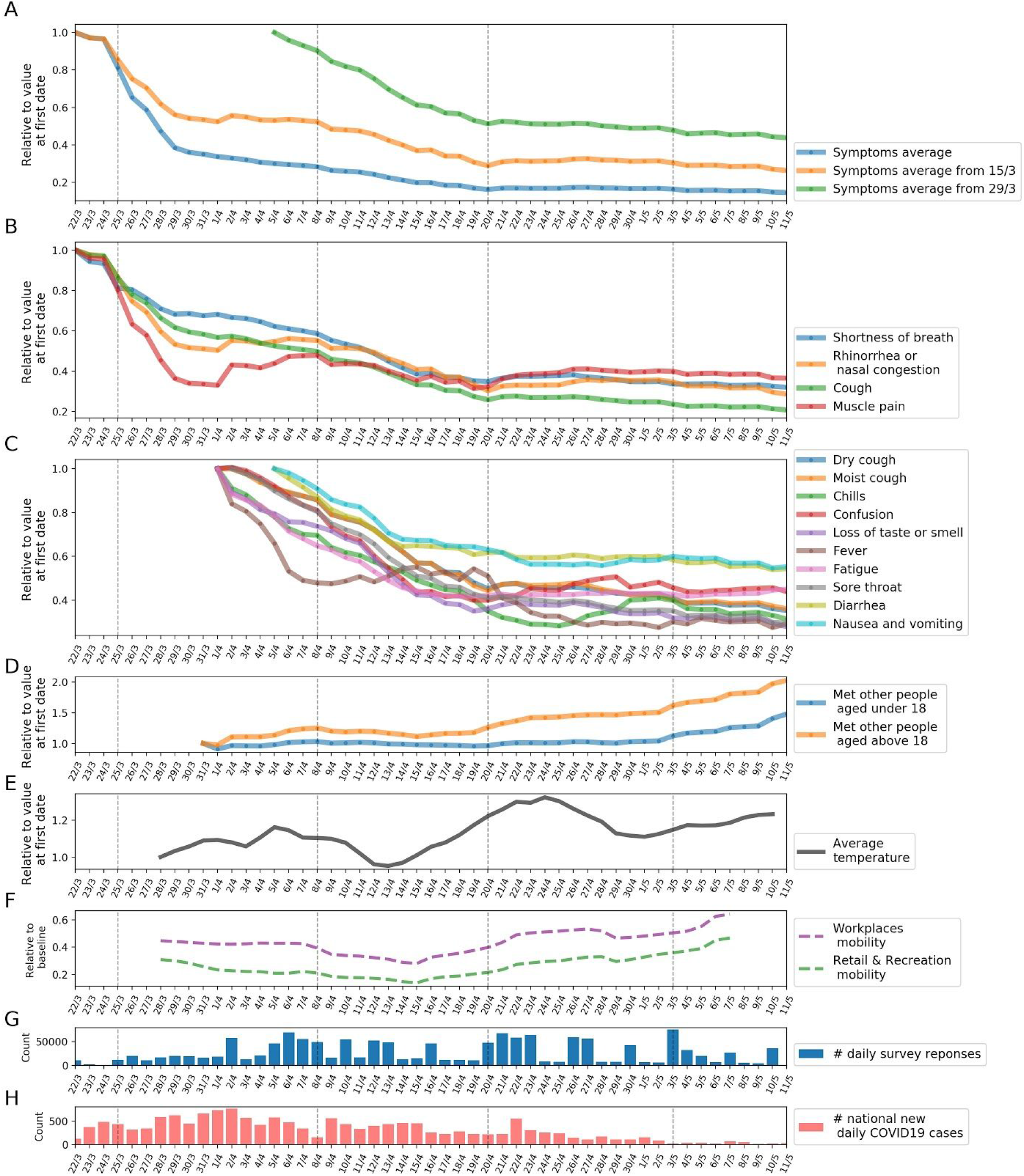
Symptoms prevalence changes from March 15th until May 11th 2020, for the full study population. **A:** Relative change of the Symptoms Average (SA) based on a 7-day running average. **B:** Relative change of symptoms reported since March 15th based on a 7-day running average **C:** Relative change of symptoms reported since March 29th based on a 7-day running average. **D:** Relative change in number of contacted people since March 15th based on a 7-day running average. **E:** Relative change in temperature since March 15th based on a 7-day running average. **F:** Relative change in mobility since March 15th based on a 7-day running average. **G:** Number of responses per day. **H:** Cumulative number of confirmed COVID-19 patients in Israel per day. **Dashed vertical lines** mark dates with COVID-19 National related events, which are shown in Table 2 (marked in bold).

Next, we calculated the prevalence of the various clinical symptoms reported by the responders throughout the study period, and analyzed the changes following policies which were implemented during this time period (Figure 1A-C, Table 2). Our analysis revealed a significant reduction in the prevalence of all symptoms as physical distancing policies became stricter. The largest reduction was observed in cough, which decreased by 74.1% from March 15th to April 20th. The most common symptom reported by responders, rhinorrhea and\or nasal congestion, decreased by 69.63%. We also observed a reduction of 83.83% in SA.

Fitting individual symptoms and the SA with a sigmoid function, we observed different slopes and shifts in the symptom’s reduction function (see Section 3 in Supplementary appendix). The slope of the function determines the rate of decrease, while the shift determines how many days after March 18th the symptom prevalence begins to decrease. The largest slope (0.509) was observed when fitting muscle pain, while the fit of shortness of breath had a much smaller slope coefficient (0.1). Some symptoms, such as moist cough and sore throat, started declining with a shift of almost 16 days from March 18th, while others like muscle pain and rhinorrhea or nasal congestion started declining earlier, with a shift of about 1.5-2 days from March 18th. These variations suggest differences in the nature of the different symptoms, which may represent different medical conditions.

Several sensitivity analyses showed a similar trend in symptoms reduction when stratifying the population by different characteristics. Geospatial analysis revealed a similar trend when separately analyzing the SA in different cities in Israel (a few sample cities are shown in Supplementary Figure S5 in Section 4 of Supplementary appendix). The decrease in the SA is also evident in all cities, despite significant population and geographical differences between them. Moreover, this trend is also visible when subgrouping the population by different demographic characteristics such as population density in the city of residence, district of residence, average number of people per household and average employment percentage, and when restricting analysis only to the subpopulation of individuals who reported at least one symptom (Supplementary Figure S6 in Section 4 of Supplementary appendix). Finally, weighting each individual in the study population using IPW, to reduce possible selection bias, also showed a similar decrease in prevalence of symptoms and SA (Supplementary Figure S2 in Section 2 of Supplementary appendix).

Following the decrease in the prevalence of individual symptoms and in the SA during the period in which physical distancing in Israel was gradually implemented, we observe a relatively stable state in the period starting from April 20th up to May 11th, when restrictions were slowly relieved. While there are slight variations in the prevalence of reported symptoms, during this time period the SA decreased only by 10%, and the most common symptom, rhinorrhea and\or nasal congestion, decreased only by 5.64% between April 20th to May 11th. Some symptoms increased in prevalence, for example fatigue and confusion which increased by 6.74% and 9.78%, respectively, from April 20th to May 11th.

Notably, the average temperature in Israel during the study period was relatively stable, with an average temperature ranging from 10-22 °C. Only a slight upward trend is visible in the average temperature, which coincides with seasonal changes in Israel during the study period, and in any case does not explain both the decrease and the final steady state that we observed in the symptoms during the study period. In addition, data obtained from the Israeli MOH on Influenza-like symptoms in Israel from previous years revealed a very low rate of influenza-like illness in the time period which is parallel to the time in which our survey has been distributed. During this time period in previous years, there were less than 1 weekly clinic visits due to influenza-like symptoms per 10,000 people, across all age groups ^11^, indicating that the symptoms reduction observed is not due to seasonal changes. Finally, although the rate of responses to the survey changes over time (Figure 1), at least 4,500 responses were received every day and a median of 16,000 responses was recorded throughout the whole study period. Thus, the decrease in all reported symptoms is probably not a result of lacking reports, but rather a true trend of reduction in symptoms.

## Discussion

Here, to the best of our knowledge, we quantify for the first time the effect of lockdown policies on the prevalence of self-reported clinical symptoms in the population. We show that implementation of various measures of lockdown across time, which manifested in decreased physical contact between individuals in the population, are accompanied by a dramatic decrease in the prevalence of numerous clinical symptoms. When these lockdown restrictions were loosened, the prevalence of these clinical symptoms remained relatively stable.

While physical distancing has been used effectively in past epidemics, decreasing human-to-human transmission and reducing morbidity and mortality ^12-14^, one of the concerns in implementing this strategy is that viral spread will be renewed upon relaxation of these measures ^13,14^. We present a data driven approach to estimate the effect of implementing physical distancing restrictions, as well as the effect of relieving them, on symptom prevalence. This may be viewed as a *Symptomete*r, which detects sensitive changes in the prevalence of symptoms in the population. Recently, we established together with other researchers an international Coronavirus Census Collective, the CCC, aimed at utilizing information on daily self-reported symptoms as means to track disease spread and predict outbreak locations ^15^. Our approach may be further implemented globally to estimate and compare different lockdown strategies taken by different countries.

Previous studies during the current COVID-19 pandemic have estimated the effects of lockdown policies on clinical variables that are more specific to COVID-19 infection, such as the effect of the lockdown on the virus doubling time ^1^. We believe that our study provides a more global view on the overall reduction in the prevalence of infectious diseases during the lockdown, including, but not limited to, COVID-19 infection. Since the symptoms included in our survey are not specific to COVID-19 infection, the prevalence of other infectious diseases is most likely reflected by them. We observe variations in the rate of decrease of different symptoms which suggests differences in the nature of different symptoms, or differences in the medical conditions they represent. This is supported by the fact that the decrease in the prevalence of symptoms is global, and is not limited to specific symptoms which were described as common in individuals with COVID-19 infection. In addition, while the number of confirmed patients with COVID-19 infection in the Israeli population is increasing (16,506 on May 12th), it still represents a small fraction (0.18%) of the Israeli population ^8^ and is unlikely to be the sole contributor to the large decrease in symptoms visible in our data. Many infectious diseases other than COVID-19 are transmitted by Aerosol transmission ^16^ or physical contact ^17^, which are greatly reduced when implementing physical distance measures, and will be therefore affected by these measures. For example, a previous study found a decrease of 42% in the overall diagnoses of respiratory infections in children during school closure as part of an organized labor dispute during a previous influenza outbreak ^18^.

One may argue that the reduction in symptoms viewed in our data is due to other factors, which are not related to the lockdown policies, such as the normal seasonal variation of Influenza infection. We believe that this is not the case, since we started distributing our survey in the middle of March, when according to data obtained from the Israeli Ministry of Health, the levels of infection caused by Influenza were already extremely low. Furthermore, data on Influenza like symptoms in Israel from previous years suggest a very low rate of influenza like illness in the time of year in which our survey was distributed (with less than 1 weekly clinic visit due to influenza like symptoms per 10,000 people) ^11^. In addition, it is unlikely to be the result of changes in the weather during this time period, as the average temperature in Israel during the study period was relatively stable. Trends in the data which directly reflect the policies enforced during the study period, such as the number of self-reported contacted people, also supports our conclusion that the observed reduction in symptoms is related to the lockdown policies.

Our study has several limitations. First, we ask participants to fill the survey anonymously since we are obligated to ensure the privacy of our participants. As such, we cannot link daily surveys of the same responder, which could have provided trends in symptoms at the individual level and insights on the progression of symptoms and the disease over time, and the influence of lockdown policies on the progression of symptoms at the individual level. At the time of writing, we are deploying newer versions of our survey that will be distributed nationally in Israel, allowing us not only to collect data which will be more representative but also to link responses of an individual over time, while protecting the anonymity and privacy of the responders.

Second, our surveys were deployed as a voluntary tool, and thus the population of the study, which was defined by the responders to the survey, is prone to selection bias, and may not represent the entire Israeli population across all geographic locations. In an attempt to reduce this bias, the survey was distributed in six different languages to reflect the most common languages spoken in Israel and substantial efforts were made to reach disadvantaged populations by engaging leaders in local religious communities, and promoting the survey through both Hebrew and Arabic-speaking media channels. Furthermore, we perform several sensitivity analyses to evaluate the robustness of our estimates, which all show similar results (Figure S5). Of note, even if selection bias exists, it will most likely be present throughout the entire study period, and is therefore less likely to lead to the relative changes that are seen in the symptom’s prevalence during the study period.

Despite the preliminary nature of our results, we believe that it is important to publish them now, as time is crucial. Overall, our study quantifies the effect of lockdown policy on the prevalence of several clinical symptoms, which characterize both COVID-19 and other infectious diseases. Such information is critical at this time, and may inform decision-makers worldwide as they consider actions to prevent the ongoing spread of coronavirus.

### Data availability statement

Tables of de-identified, aggregated data are available at https://github.com/hrossman/Covid19-Survey.

### Code availability statement

Analysis source code is available at https://github.com/hrossman/Covid19-Survey. Source code for the questionnaires is available at https://github.com/hasadna/avid-covider as an open source project, and can be readily adapted to use in other countries.

## Ethics Declarations

The study protocol was approved by the Institutional Review Board (IRB) of the Weizmann Institute of Science. Informed consent was waived by the IRB, as all identifying details of the participants were removed before the computational analysis. Participants were made fully aware of the way in which the data will be stored, handled and shared, which was provided to them and is in accord with the privacy and data-protection policy of the Weizmann Institute of Science (https://weizmann.ac.il/pages/privacy-policy).

## Authors contribution

A.K., A.G., H.R., S.S. & T.M. conceived the project, designed and conducted the analyses, interpreted the results and wrote the manuscript. T.K., A.L., D.K., I.K., S.S., & N.G. designed and conducted the analyses, interpreted the results and wrote the manuscript. O.C. directed the organizational and logistic effort and interpreted the results. A.A.Z & A.I. provided and interpreted data. B.G, Y.D., R.B., V.S. & E.S. conceived and directed the project and analyses, designed the analyses, interpreted the results, wrote the manuscript, and supervised the project.

## Acknowledgments

We thank the following for their contributions to our efforts: Y. Landau, T. Eldar, S. Kasem, T. Bria, S. Avraham, B. Kirel, A. Terkeltaub, N. Kalkstein, E. Krupka, M. Loitsker, Y. Sela, T. Bareia, J. Shalev, A. Brik, L. Samama, S. Cohen, J. Bitton, R. Priborkin, G. Beryozkin, G. Dardyk, S. Lanzman, N. Epstein, N. B. Shizaf, E. Daian, M. Bialek, M. Suleiman, R. Maor, Z. Maor, Y. Dinur, J. Klinger, R. Zaida, M. Eden, N. Hirshman, O. Dovev, Y. Grossman and the Public Knowledge Workshop (‘Hasadna’).

## Supplementary appendix

1. COVID-19 daily questionnaire
2. Inverse Probability Weighting
3. Modelling symptoms dynamics
4. Sensitivity Analysis

## 1. COVID-19 daily questionnaire

Integration time of different symptoms to our survey:

**Supplementary Figure S1:**
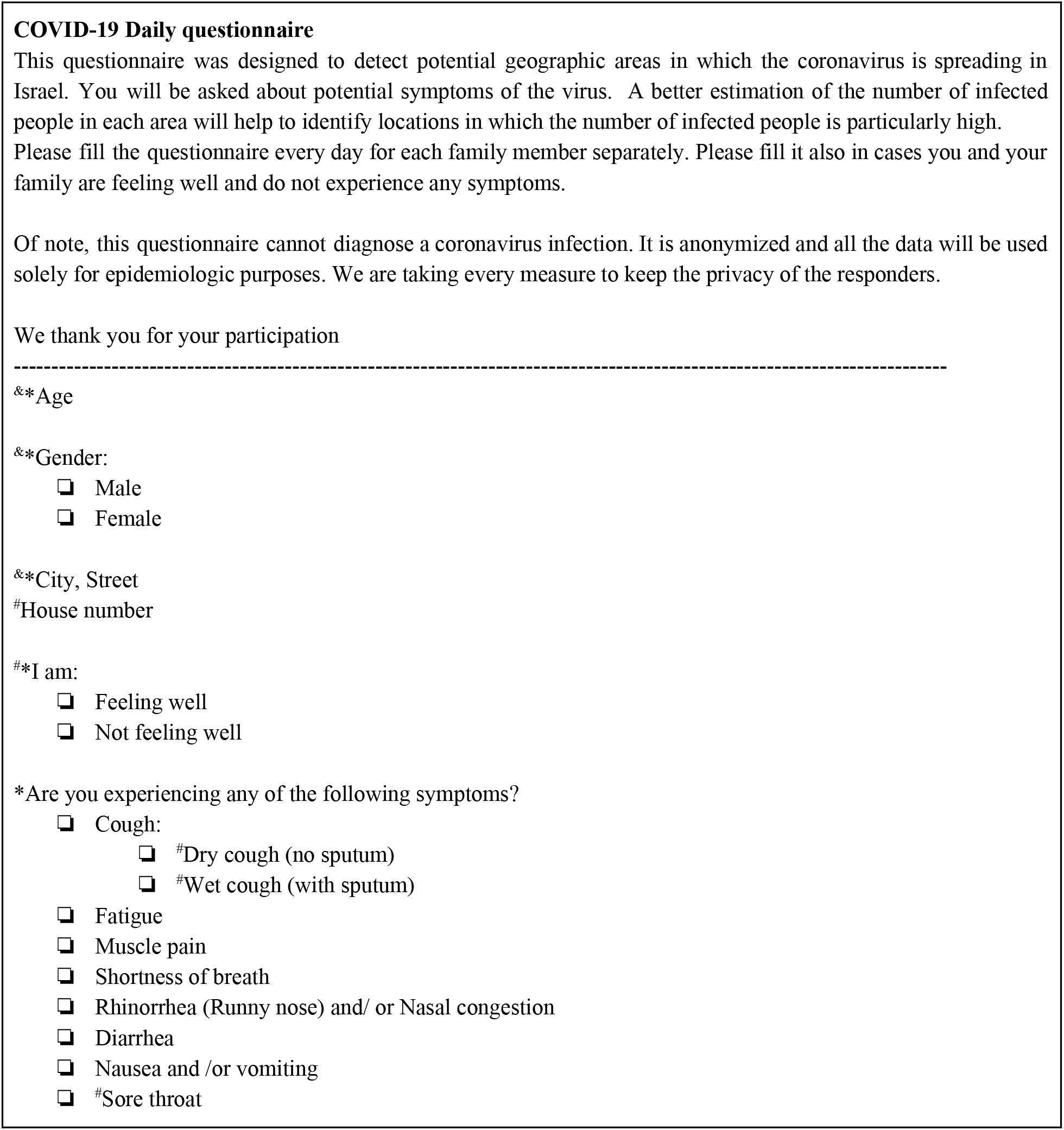
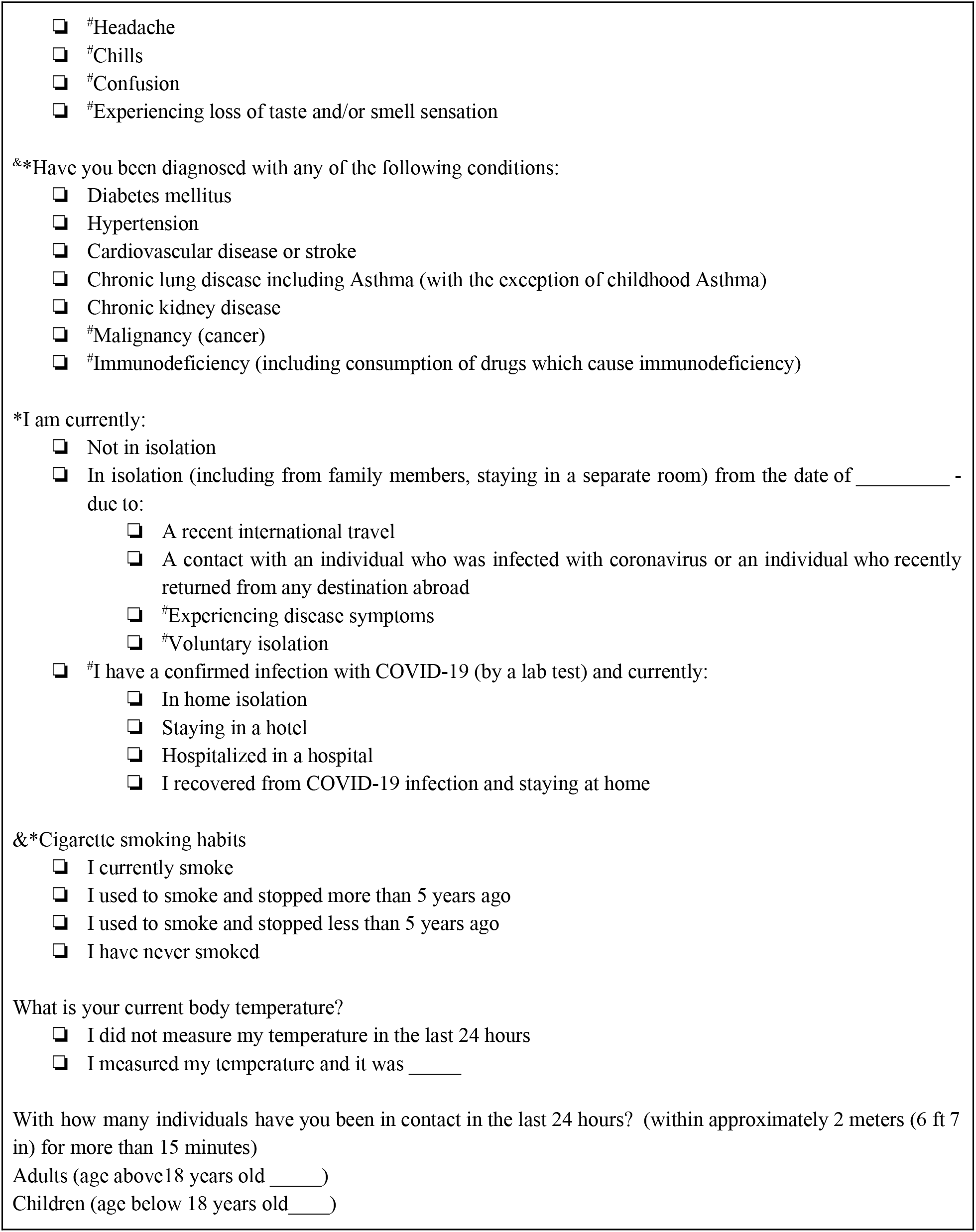
COVID-19 Daily questionnaire *Questions that the responder is required to answer, & Questions that should be filled only once #Questions that were added in newer versions of the questionnaire

- Symptoms reported from March 15th: shortness of breath, rhinorrhea or nasal congestion, cough, fatigue, nausea and vomiting, muscle pain, fever, diarrhea. We define SA from 15/3 as the average of these symptoms.
- Symptoms reported from March 29th: cough, chills, confusion, loss of taste or smell, sore throat. We define SA from 29/3 as the average of these symptoms.

## 2. Inverse Probability Weighting

As our survey is voluntary, the population of responders may suffer selection bias. To estimate the sensitivity of our results to this bias we employed weighting of the individual data so it will better represent the true target population (Israel’s population), thus reducing selection bias. Weights are calculated by binning individuals into city and age group bins, and then weighting each bin to represent its true proportion in the target population. City was defined as a geographical area as defined by the Israel Central Bureau of Statistics (ICBS) ^20^. Respondents were associated with cities using the address provided in the questionnaire. Age was divided into 4 groups; 0-17, 18-34, 35-54 and 75-120, using the ICBS information, and each responder was assigned an age bin using the age provided in the survey.

**Supplementary Figure S2:**
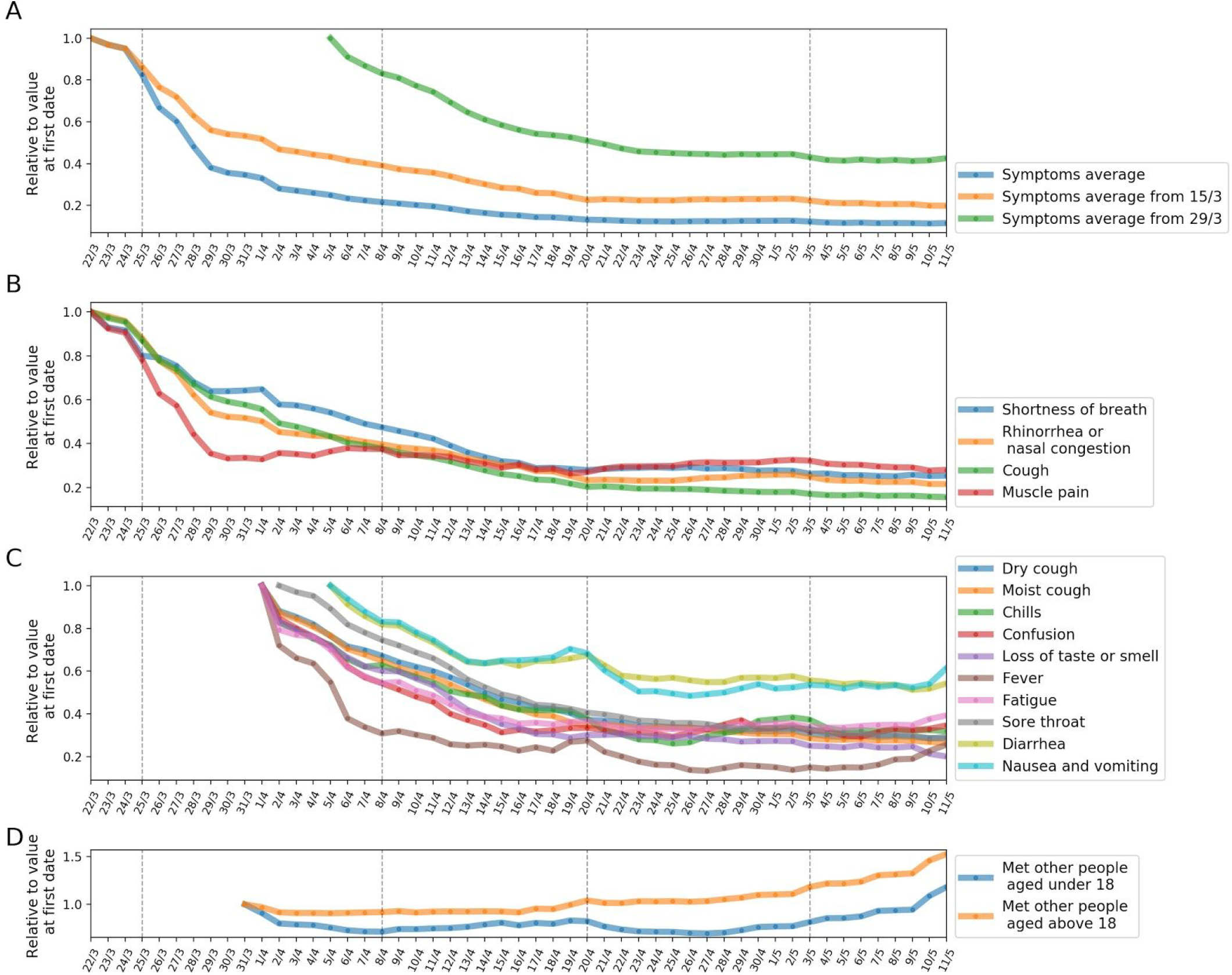
Symptoms prevalence changes from March 15th until May 11th 2020, for weighted study population. **A:** Relative change of the Symptoms Average (SA) based on a 7-day running average. **B:** Relative change of symptoms reported since March 15th based on a 7-day running average **C:** Relative change of symptoms reported since March 29th based on a 7-day running average. **D:** Relative change in number of contacted people since March 15th based on a 7-day running average. **Dashed vertical lines** mark dates with COVID-19 National related events, which are shown in Table 2 (marked in bold).

## 3. Modelling symptom dynamics

We define the function as follows:

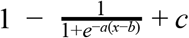

Where x is the aggregated mean of the symptom over the population, and a, b, c are the coefficients we fit. *a* determines the slope of the function, the bigger the value the steeper the slope. *b* centers the function and determines where the slope starts on the *x* axis, the bigger it is, the later in time the function will decrease. *c* determines where the slope starts on the *y* axis, the bigger it is the higher the symptom average was in time zero.

**Supplementary Table S1:** Coefficients of fitted sigmoid functions on symptoms. The table presents the fitted coefficients of the sigmoid function. *a* determines the slope of the function, *b* centers the function and determines where the slope starts on the *x* axis. *c* determines where the slope starts on the *y* axis

| Symptom | a (slope) | b (days since 18/3) | c (base rate) |
| --- | --- | --- | --- |
| Muscle pain | 0.509724538 | 1.785851231 | 0.388448392 |
| Chills | 0.142011429 | 14.47764421 | 0.211289502 |
| Diarrhea | 0.215262878 | 12.17349717 | 0.561035886 |
| Rhinorrhea or nasal congestion | 0.173724971 | 1.540542654 | 0.383806857 |
| Fatigue | 0.204151026 | 11.0935456 | 0.377927786 |
| Fever | 0.370279661 | 9.98679371 | 0.418760017 |
| Cough | 0.132664781 | 5.32907753 | 0.263384775 |
| Nausea and vomiting | 0.154839032 | 13.05960585 | 0.499199853 |
| Loss of taste or smell | 0.156707564 | 13.6565442 | 0.287932174 |
| Confusion | 0.177337307 | 13.8026112 | 0.340404527 |
| Moist cough | 0.126542255 | 15.91251757 | 0.32863404 |
| Dry cough | 0.125325423 | 16.39732625 | 0.314183738 |
| Sore throat | 0.156284544 | 15.51714757 | 0.305211429 |
| Shortness of breath | 0.099004087 | 5.261930633 | 0.297520094 |
| Symptoms average | 0.372379649 | 3.774412671 | 0.217189425 |
| Symptoms average from March 15th | 0.15047274 | 2.650949218 | 0.336042214 |
| Symptoms average from March 29th | 0.169276153 | 14.92601557 | 0.432761618 |

**Supplementary Figure S3:**
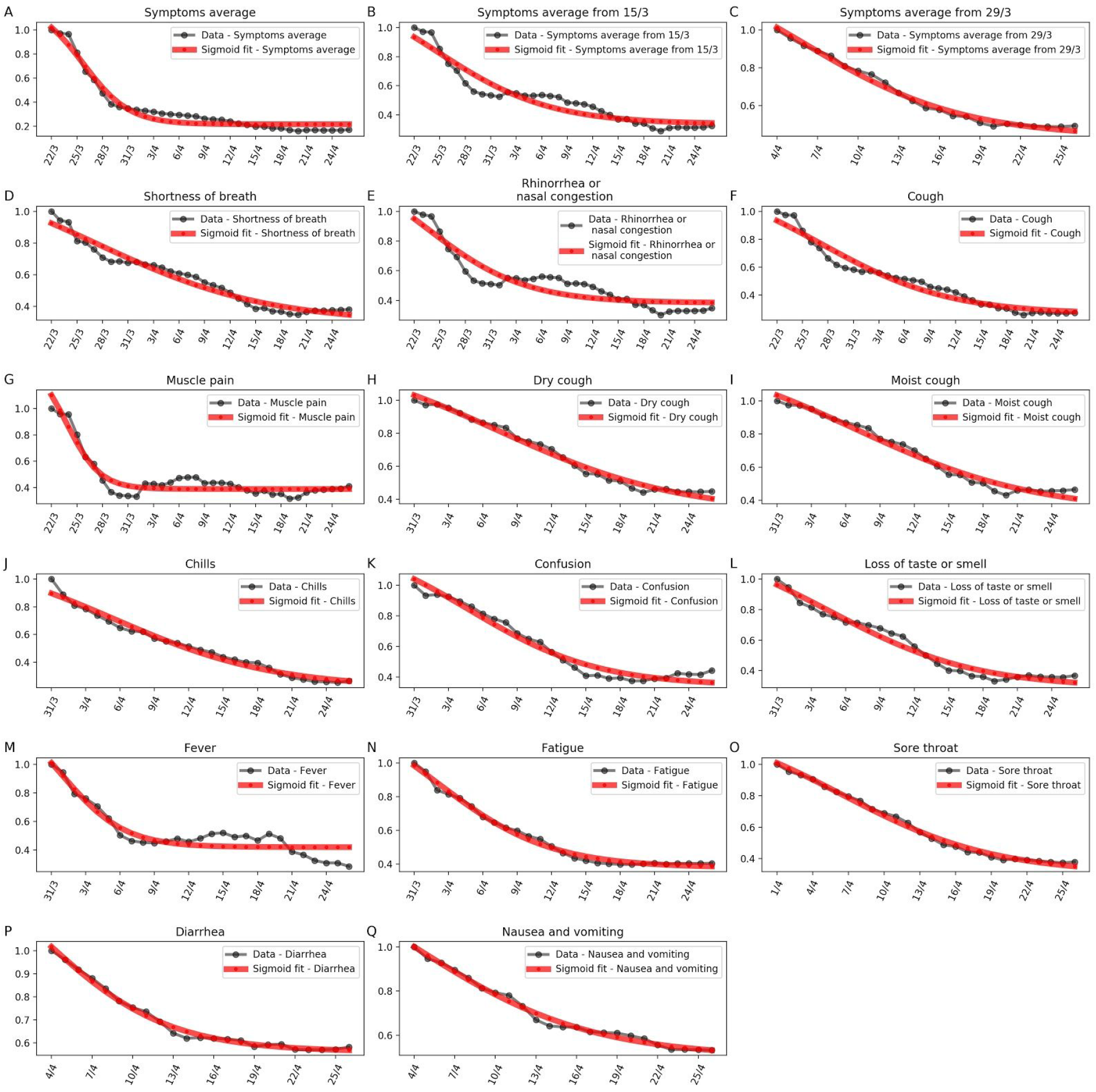
Modeling symptoms dynamics. Each panel shows the relative change in symptoms prevalence since its integration time to our survey. Black dots - actual relative value, red line - fitted function.

**Supplementary Figure S4:**
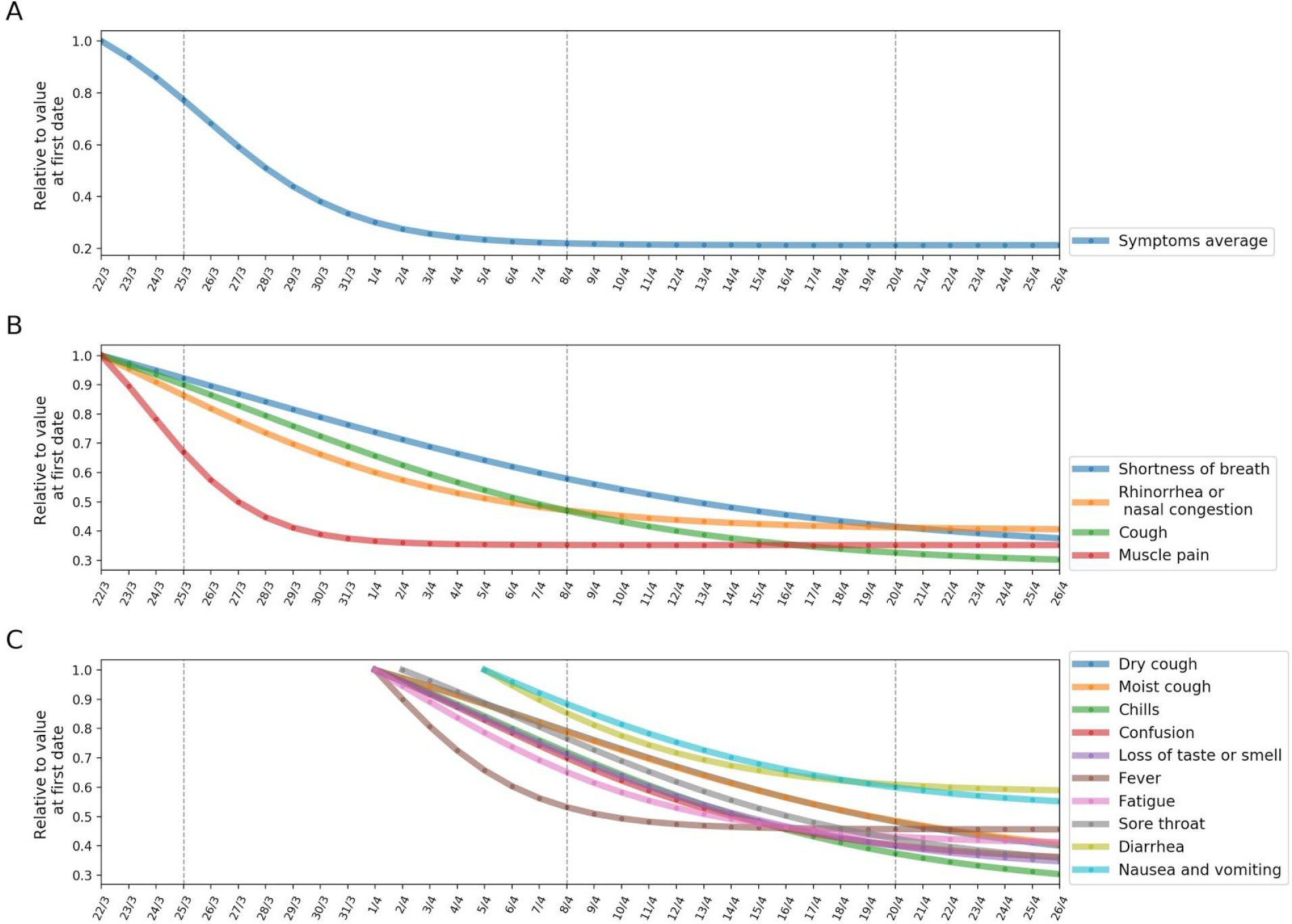
Fitted sigmoid function of symptoms decrease from March 18th until April 22nd, 2020 in study population. **A:** Different symptoms decrease over time. In each subplot the black curve is the 3-day running average and the red curve is the fitted sigmoid function over the symptom. **B:** Relative decrease of the fitted sigmoid function SA based on a 3-day running average. **C:** Relative decrease of the fitted sigmoid function on different symptoms which were collected prior to the 18th of April based on a 3-day running average. **D:** Relative decrease of the fitted sigmoid function on different symptoms which were collected from the 24th of April based on a 3-day running average.

## 4. Sensitivity Analysis

Stratification of the population by several demographic characteristics was done by associating each response with a residence city, according to the address reported in the survey. Cities’ demographics features such as population density, and average housing density were obtained from the Israel Central Bureau of Statistics. An additional subpopulation of symptomatic individuals was defined by the responses from the survey which contain at least one reported symptom.

**Supplementary Figure S5:**
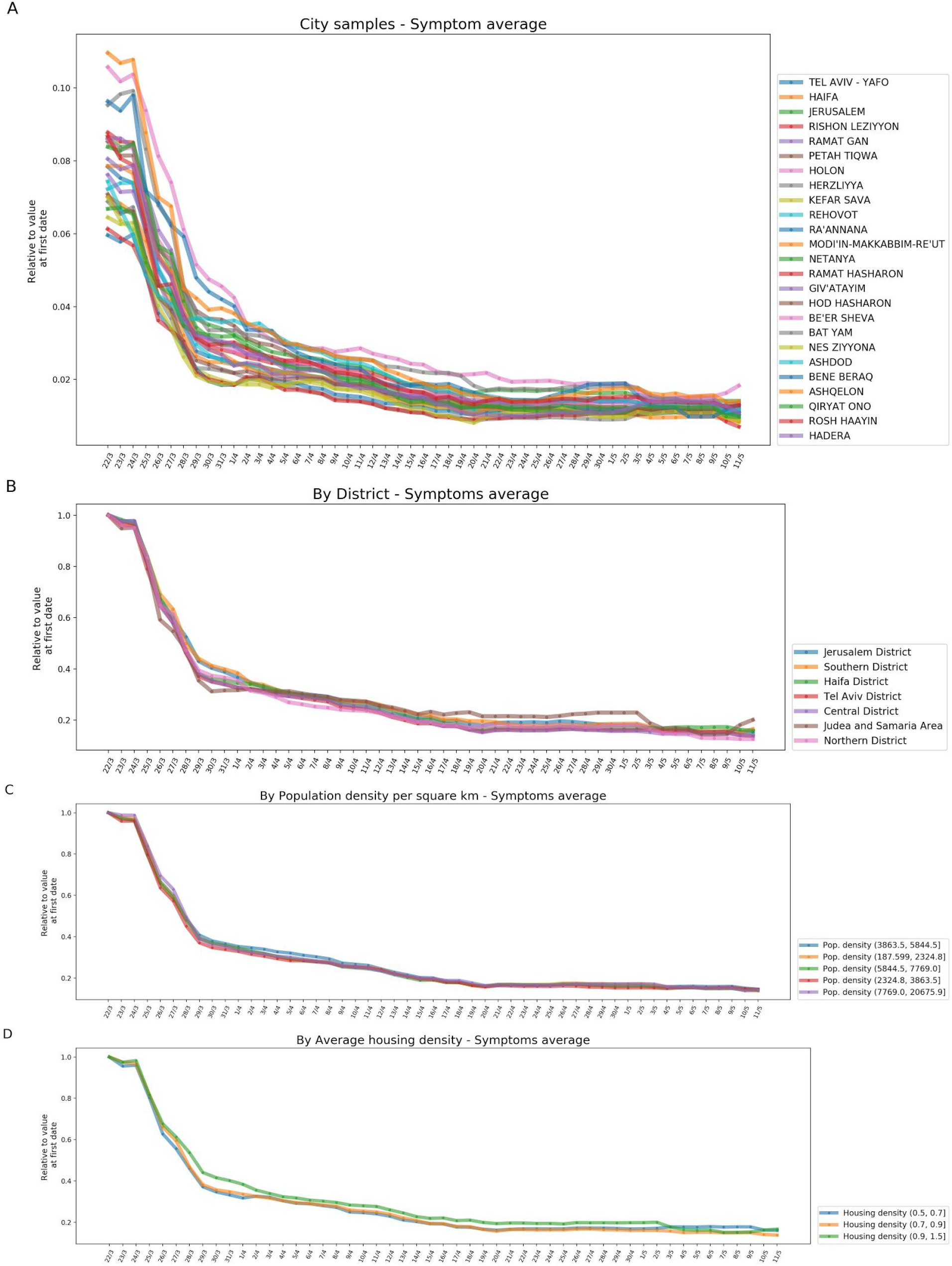
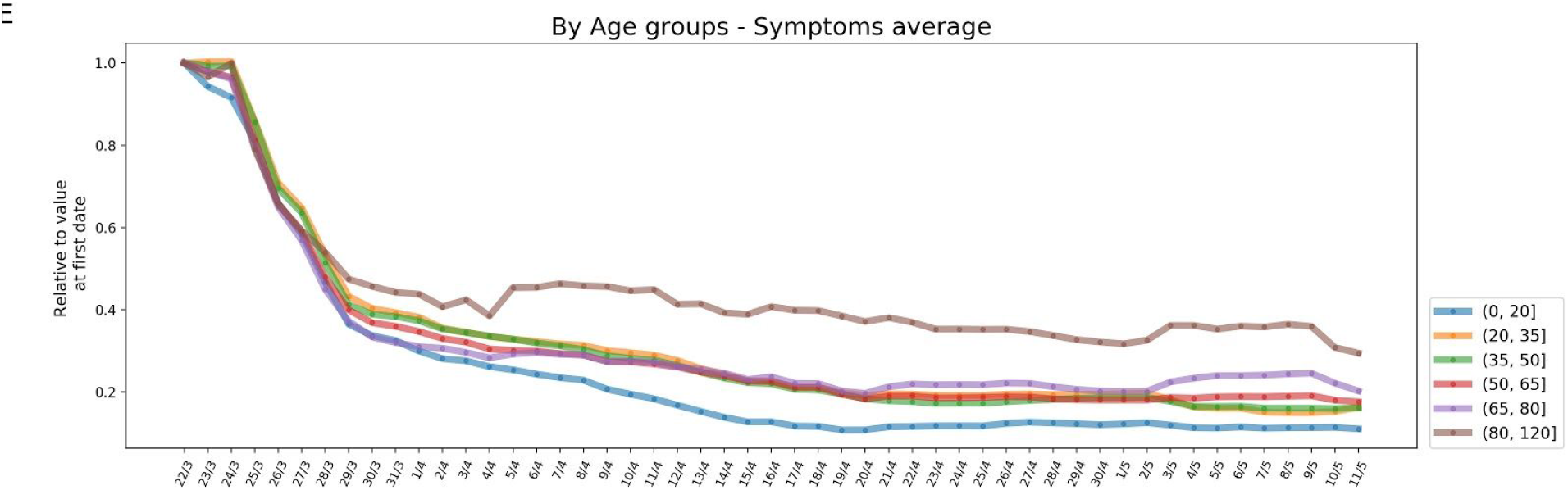
Symptoms Average Relative values from March 15th until May 11th, 2020 in study population subgroups. **A:** Selected cities. **B:** Geographic districts. **C:** Subgroups by residence city’s population density. **D:** Subgroups by average housing density in residence city. **E:** Subgroups by age groups.

**Supplementary Figure S6:**
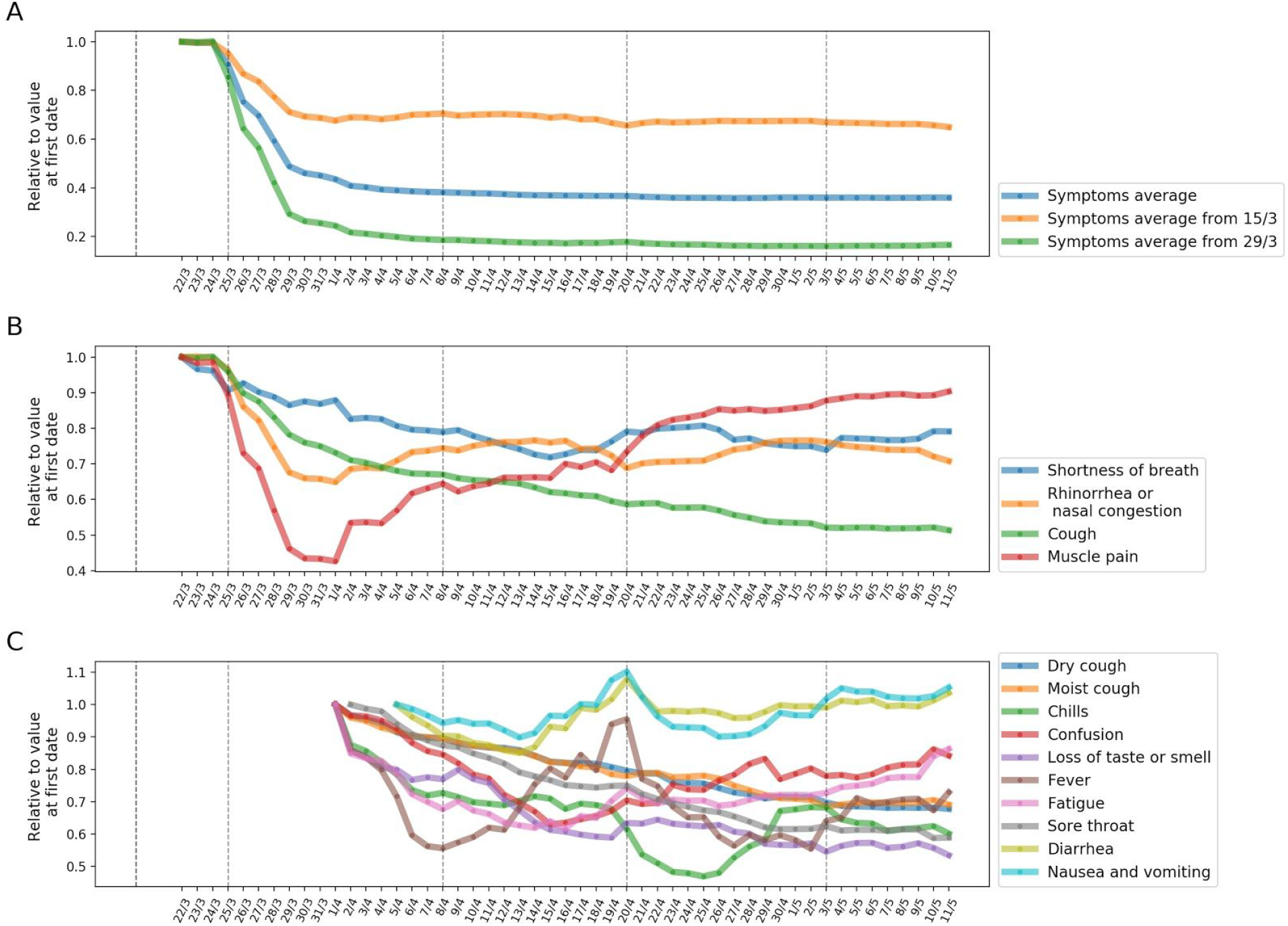
Symptoms prevalence changes from March 15th until May 11th 2020, in subpopulation of symptomatic. **A:** Relative change of the Symptoms Average (SA) based on a 7-day running average. **B:** Relative change of symptoms reported since March 15th based on a 7-day running average **C:** Relative change of symptoms reported since March 29th based on a 7-day running average.

**Supplementary Figure S7:**
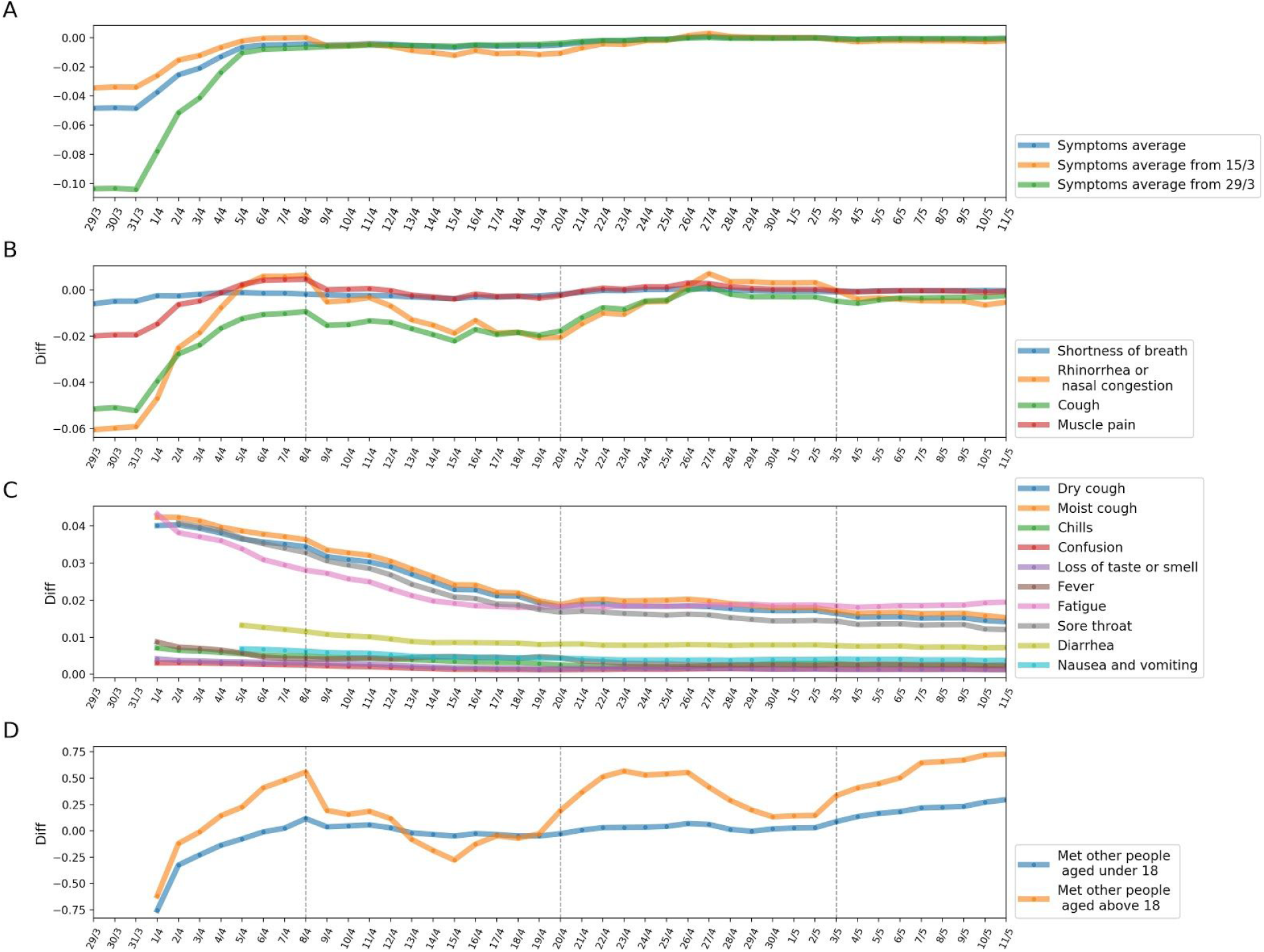
Symptoms prevalence 7-day differences from March 15th until May 11th 2020. **A:** 7-day difference of the Symptoms Average (SA) based on a 7-day running average. **B:** 7-day difference of the symptoms reported since March 15th based on a 7-day running average **C:** 7-day difference of symptoms reported since March 29th based on a 7-day running average. **D:** 7-day difference in number of contacted people since March 15th based on a 7-day running average.

